# *HSP90AA1* variants may contribute to autosomal dominant human male infertility

**DOI:** 10.64898/2026.01.08.26343516

**Authors:** Margot J. Wyrwoll, David MacKenzie MacLeod, Ahmet Salvarci, Sabine Kliesch, Özlem Okutman, Stéphane Viville, Birgit Stallmeyer, Frank Tüttelmann, Dónal O’Carroll

## Abstract

**Study question:** Do variants in *HSP90AA1* cause human male infertility?

**Summary answer:** Variants in *HSP90AA1* appear as a possible autosomal dominant cause of human male infertility.

**What is known already:** Male infertility is a highly heterogeneous condition, with so far over 300 genes described in this context. *HSP90AA1* appears as a promising candidate gene for human male infertility, because the gene is highly conserved between species and knock-out of *Hsp90aa1* in mice results in male-specific infertility due to azoospermia without further health implications.

**Study design, size, duration:** We screened >2,300 infertile men for possibly pathogenic variants in *HSP90AA1* and created a mouse line harbouring the homozygous missense variant c.605G>A p.(Arg202Lys).

**Participants/materials, setting, methods:** Phenotypes of men with identified variants were determined based on semen analysis and testicular histology. Pathogenicity of detected variants was assessed using AlphaMissense and a mouse model. Male fertility of the mutant mouse line was analysed via plug-matings, histology and immunofluorescence staining (IF). Expression of HSP90AA1 in testicular tissue was assessed by IF.

**Main results and the role of chance:** The mode of inheritance (MOI) in mice is autosomal recessive but the constraint metrics (oe-score = 0.2, pLI = 1) and *in silico* prediction suggest that *HSP90AA1* is an autosomal dominant gene in humans. We therefore screened for both, heterozygous and biallelic variants in exome sequencing data of infertile men. While we did not detect any biallelic loss-of-function variants, we identified the homozygous missense variant c.605G>A p.(Arg202Lys) in an azoospermic man as a promising variant. This variant is extremely rare and affects a highly conserved amino acid. However, male homozygous mice with this variant are fertile with no differences in litter size and testicular size or histology, making it unlikely that this variant is the cause of the man’s azoospermia. We therefore focused on heterozygous possibly pathogenic variants in *HSP90AA1* and found a heterozygous frameshift variant in an azoospermic man with hypospermatogenesis as well as four heterozygous missense variants, predicted to affect protein function in azoo- or cryptozoospermic men.

**Large scale data:** N/A

**Limitations, reasons for caution:** Our findings suggest a dominant MOI in humans but currently cannot fully prove this. To further clarify the MOI and ultimately improve clinical validity of *HSP90AA1* replication of our findings in independent cohorts of infertile men as well as segregation analyses are required.

**Wider implications of the findings:** While most human male infertility genes follow an autosomal recessive MOI, *HSP90AA1* might be one of the few autosomal dominant infertility genes in humans. Differences in the MOI between humans and mice are also known from well-established infertility genes such as *DMRT1*.

**Study funding/competing interest(s):** This work was supported by a German Research Foundation (DFG) fellowship (award WY 215/1-1 to MJW), the DFG-sponsored Clinical Research Unit ‘Male Germ Cells’ (CRU326, project 329621271 to FT), and Wellcome Trust funding (225237 to DOC). This work was supported by funding for the Wellcome Discovery Research Platform for Hidden Cell Biology (226791).

## Introduction

Infertility represents a major public health issue with around 15% of all couples worldwide being affected (WHO, 2023). In around half of the cases, a male factor can be identified as the underlying cause for the couple’s infertility. While there are many non-genetic causes of male infertility such as testicular cancer or previous chemotherapy, it has previously been shown that a large fraction of infertility cases is due to genetic alterations (Wyrwoll *et al*., 2023). Azoospermia represents the most severe male infertility phenotype with no sperm detected in the semen. It is assumed to be of genetic origin in many cases (James *et al*., 2025). In the last ten years numerous monogenic causes of human azoospermia were revealed, increasing the monogenic diagnostic yield to around 6% (Riera-Escamilla *et al*., 2025). Detection of a genetic cause of azoospermia can help to predict the chances of successful testicular sperm extraction (TESE) in azoospermic men (Wyrwoll *et al*., 2023; Riera-Escamilla *et al*., 2025). However, most affected men remain without a causal diagnosis. Deciphering the genetic causes of human azoospermia will not only help to predict the chances of TESE and provide a causal diagnosis for affected men but also improve our understanding of spermatogenesis which might ultimately pave the way for novel treatment options.

Discovery of many genes linked to human azoospermia was facilitated by sequencing genes in infertile men, which have previously been described to result in azoospermia in mice when genetically altered (Riera-Escamilla *et al*., 2019). As the process of gametogenesis and especially meiosis is highly conserved in evolution (Brattig-Correia *et al*., 2024), the mouse model represents an adequate model for most infertility genes. However, the mode of inheritance may vary between mice and humans (James *et al*., 2025) as e.g. known for variants in *DMRT1*, which follow and autosomal recessive mode of inheritance in mice (Raymond *et al*., 2000) and an autosomal dominant mode of inheritance in humans (Emich *et al*., 2023).

A gene that is linked to azoospermia in mice is *Hsp90aa1*. Biallelic knock-out (KO) of *Hsp90aa1* leads to male specific infertility in mice without further malformations (Grad *et al*., 2010). Homozygous male *Hsp90aa1*^*-/-*^ mice exhibit azoospermia due to spermatocyte arrest at pachytene stage of meiosis 1, while heterozygous male *Hsp90aa1*^*+/-*^ mice have normal fertility (Grad *et al*., 2010). *Hsp90aa1* encodes the heat shock protein 90 (HSP90) paralogue HSP90α, which is highly similar to the isoform HSP90 paralogue HSP90β, encoded by *Hsp90ab1*. HSP90α is primarily expressed in the testis (Grad *et al*., 2010) whereas HSP90β is ubiquitously expressed (Sreedhar *et al*., 2004). *Hsp90ab1*^*-/-*^ mice are not viable due to malformations of the placenta (Voss *et al*., 2000). HSP90AA1 is a highly conserved protein with >99% sequence identity between humans and mice. It has three conserved domains, an N-terminal ATP-ase domain, a middle domain mediating client binding and a C-terminal dimerization domain (Zuehlke *et al*., 2015). So far, no pathogenic variants have been reported in *HSP90AA1*, although single nucleotide polymorphisms (SNPs) in this gene have been associated with human male infertility (Xue *et al*., 2021). In this study we aimed at investigating the role of *HSP90AA1* as a novel candidate gene for human male infertility.

## Materials and Methods

### Ethical approval

The study protocol was approved by the respective local ethics committees: MERGE cohort Münster (2010-578-f-S), Strasbourg (CPP 09/40—WAC-2008-438 1W DC-2009-I 002), and Yeni Yüzyıl University, scientific, social and noninterventional health sciences research ethics committee, Istanbul, Turkey (approval no: 2019/08). All persons gave written consent compliant with local requirements and all experiments were performed in accordance with the criteria set by the Declaration of Helsinki.

### Study cohort

At the time of the query, the MERGE cohort included exome/genome sequencing data of 2,386 men in infertile couples with various phenotypes. Most of these men were recruited at the Centre of Reproductive Medicine and Andrology (CeRA) in Münster and are azoospermic (HPO:0000027; N = 1,448), cryptozoospermic (HPO:0030974; N = 454), extreme oligozoospermic (HPO:0034815; N = 158), or severe oligozoospermic (HPO:0034818; N=67). Twenty-three of the aforementioned men with azoospermia were recruited in Strasbourg including men from Istanbul.

### Mode of inheritance predictions

To determine the mode of inheritance for *HSP90AA1* in humans the constraint metrics provided by gnomAD (Karczewski *et al*., 2020), the pHaplo (Collins *et al*., 2022) and the prediction by DOMINO (Quinodoz *et al*., 2017) were taken into account.

### Exome/genome sequencing

Sequencing and bioinformatics analyses in the MERGE cohort were performed as previously described (Wyrwoll *et al*., 2023). In brief, genomic DNA was extracted from peripheral blood leukocytes via standard methods. Sample preparation and enrichment for exome sequencing was carried out according to the protocol of either Agilent’s SureSelectQXT Target Enrichment for Illumina Multiplexed Sequencing Featuring Transposase-Based Library Prep Technology (Agilent) or Twist Bioscience’s Twist Human Core Exome. For genome sequencing, libraries were prepared according to Illumina’s DNA PCR-Free library kit. Sequencing was carried out on Illumina HiSeq 4000/X, NextSeq 500/550, or NovaSeq 6000.

### Variant calling

Cutadapt v1.15 was used for trimming of remaining adaptor sequences and primers. Reads were then aligned against GRCh37.p13 using BWA Mem v0.7.17. Variant calling and base quality recalibration were performed using the GATK toolkit v3.8 (McKenna *et al*., 2010). Samples undergoing whole genome sequencing were aligned with Illumina Dragen Bio-IT platform v4.2. For both pipelines the reference genome GRCh37.7.p13 was used. Annotation of the resulting variants was done with Ensembl Variant Effect Predictor (McLaren *et al*., 2016).

### Screening of exome/genome sequencing data

We filtered the exome sequencing data for variants in *HSP90AA1* (NM_005348.4) with a minor allele frequency (MAF) ≤0.001 listed in gnomAD v2.1.1 (Karczewski *et al*., 2020), a CADD score for missense variants ≥20 and only occurring once in our in-house database. Patients with previously identified genetic causes of infertility were excluded from further analysis.

All men with *HSP90AA1* variants were additionally screened for alternative genetic variants which might explain the fertility phenotype. To this end we screened for rare (MAF ≤0.01) homozygous and hemizygous variants with a CADD score >20 or predicted loss-of-function variants.

### AlphaFold2 protein structure

AlphaFold2 structure prediction and pathogenicity assessment was obtained from https://alphamissense.hegelab.org/.

### Human histology

Testicular biopsies of patients from the MERGE cohort and control subjects were obtained from patients who underwent testicular sperm extraction (TESE) at the Department of Clinical and Surgical Andrology at the University Hospital Münster, Germany. The testicular biopsies were fixed in Bouin’s solution overnight, subsequently washed with 70% ethanol and embedded in paraffin for routine histological evaluation. Next, 5 µm sections were stained with periodic acid-Schiff (PAS) according to previously published protocols (Brinkworth *et al*., 1995). The sections were dewaxed in solvent (ProTaqs Clear, #4003011; Quartett Immunodiagnostika and Biotechnologie, Berlin, Germany) and rehydrated in a decreasing ethanol series. Afterwards they were incubated for 15 min in 1% periodic acid. Next, they were washed with dH2O and incubated for 45 min with Schiffs reagent (Roth, Karlsruhe, Germany). In the last step, the slides were cleared for 2 × 5 min in xylene and finally mounted with Pertex® mounting medium (Histolab #00801).

### Mouse strain

The *Hsp90aa1*^*R202K*^ allele was created using CRISPR-Cas9 gene editing technology. For this we used B6CBAF1/Crl genetic background fertilized 1-cell zygotes as previously described (Wang *et al*., 2013; Yang *et al*., 2013). A single sgRNA (TTTGGAGGAAAGGAGAATAA) was injected together with Cas9 mRNA in *Hsp90aa1*^*R202K*^ mutation (AGA>AAG) and a PAM site mutation flanked by 98 and 92 nucleotides of homology arms (AACCATTGCCCTTTTCTTTTCCAGGTGAACCAATGGGTCGTGGAACAAAGGTTATCTTGCATCTGAAAGAAGACCAAACAGAGTATTTGGAGGAAAGGAGAATAAAGGAGATCGTGAAGAAGCATTCTCAGTTCATTGGCTATCCCATTACTCTCTTTGTAAGTTACCTACAGGGTAAATTTCATATAATTGAGATAACT). F0 offspring were screened by PCR and Sanger sequencing to verify the sequence of the *Hsp90aa1*^*R202K*^ allele. The line was established from one founder animal and back-crossed several times to a C57BL/6N genetic background. Thus, the mice analysed were on a mixed B6CBAF1/Crl; C57BL/6N genetic background. *Hsp90aa1*^*R202K*^ mice were genotyped by PCR using the following primer pairs: F1 TGTATGGCTGTTTGCTCAGTTG, F2 CAGAGTATCTTGAGGAGCGC, R1 GCTTCTTCACGATCTCCTTTATTC, R2 CAGCCTCATCATCACTGACTTC.

Fertility of male mice was assessed by mating *Hsp90aa1*^*R202K/R202K*^ and wildtype studs to C57BL/6N wildtype females, counting the number of pups born from each plugged female. All animals were maintained at the University of Edinburgh, UK in accordance with the regulation of the UK Home Office. Ethical approval for the mouse experimentation has been given by the University of Edinburgh’s Animal Welfare and Ethical Review Body. All work was done under licence from the United Kingdom’s Home Office.

### Mouse histology

Isolated testes and epididymes of 2-months-old mice were fixed overnight in Bouin’s fluid, washed three times in 70 % ethanol and embedded in paraffin. 6 μm sections were cut on a microtome (Leica) and deparaffinised in a graded alcohol series according to standard laboratory procedures. The rehydrated sections were then stained with the PAS staining kit (TCS Biosciences) according to the manufacturer’s recommendations. The stained sections were subsequently de-hydrated in a reverse alcohol series and mounted on coverslips with Pertex mounting media (Pioneer Research Chemicals) according to standard laboratory procedures. Slides were imaged on a Zeiss AxioScan scanning microscope using the 40x objective. Cropped images of the scan were exported using the Zeiss Zen software and further processed in ImageJ.

### Immunofluorescence

For immunofluorescence 6 μm sections were freshly cut from OCT embedded testes from 2-months-old mice as previously described (Vasiliauskaitė *et al*., 2017). Primary antibodies were incubated overnight in blocking buffer (dilutions: anti-HSP90AA1 (PA3-013, Invitrogen) 1:2000, anti-SCP1 (ab15090, abcam) 1:300, anti-SCP3 (D1, sc74569, Santa Cruz Biotechnology) 1:300). Next, sections were stained with DAPI and the appropriate donkey anti-rabbit or donkey anti-mouse secondary antibodies labelled with Alexa Fluor dyes (488, 568 or 647). Glass coverslips were then mounted onto sections with Prolong Gold (Invitrogen). All images were acquired on a Zeiss Observer with a Leica SP8 confocal microscope or Zeiss LSM880 with Airyscan module. Images acquired using the Airy scan module underwent deconvolution using the Zeiss Zen software “Airyscan processing” on settings “3D” with a strength of 6. Finally, images were processed and analysed with ImageJ and the Zeiss Zen software.

## Results

### Determination of the mode of inheritance

As male homozygous *Hsp90aa1*^*-/-*^ mice but not male heterozygous *Hsp90aa1*^*+/-*^ mice are infertile, an autosomal recessive mode of inheritance for pathogenic variants in *HSP90AA1* in humans seemed likely. However, the constraint metrics in humans point towards an autosomal dominant mode of inheritance. Constraint metrics are quantitative measures of how intolerant a human gene is to genetic variation, inferred from how depleted that gene is for observed variants compared with what would be expected under neutrality in large population datasets. We found *HSP90AA1* to have an pLI score of 1. A pLI score of >0.9 indicates haploinsufficiency and is predominantly seen in genes with a dominant mode of inheritance. As constraint metric for intolerance towards Lof variants the LOUEF score is recommended to be used, because it provides a continuous value. In gnomAD4.1 a threshold of <0.6 is recommended with *HSP90AA1* having a LOEUF score of 0.24, highly indicative of an autosomal dominant mode of inheritance. According to Colins *et al. HSP90AA1* has a pHaplo of 0.96 (Collins *et al*., 2022) with a pHaplo >0.51 as a reference for high confidence haploinsufficient genes. Genes with pHaplo scores in this range contribute to higher dosage sensitivity annotations. These findings are further supported by the DOMINO prediction of the gene being “very likely dominant” (Quinodoz *et al*., 2017)(Figure 3B). Additionally, the Z-score of 2.78 suggests an increased intolerance to missense variation. Based on the contradicting information from mouse model and prediction tools, we did not decide for or against one of the inheritance modes but screened the exome sequencing data for heterozygous as well as homozygous variants.

### Homozygous *HSP90AA1* variant p.(Arg202Lys)

The only biallelic variant fulfilling our filtering criteria was the homozygous missense variant hg38/GRCh38: c.605G>A p.(Arg202Lys) with a CADD score of 24.2 and a MAF of 0.0000064 which was not found in homozygous state in any individual in gnomAD database but three times in heterozygous state. Further it was not found in any other individual in our in-house database. The affected man M3272 is azoospermic due to Sertoli cell-only (SCO) phenotype (Figure S1A) and is son of consanguineous parents (Figure S1B). This variant affects an amino acid residue that is conserved across vertebrates including as far as lamprey (Figure 1A). The variant lies in the N-terminal domain of HSP90AA1, suggesting that ATP binding or hydrolysis might be impaired by the amino acid exchange. We therefore generated a mouse line carrying this variant to investigate its potential pathogenicity (Figure S1C, D, E).

**Figure 1.**
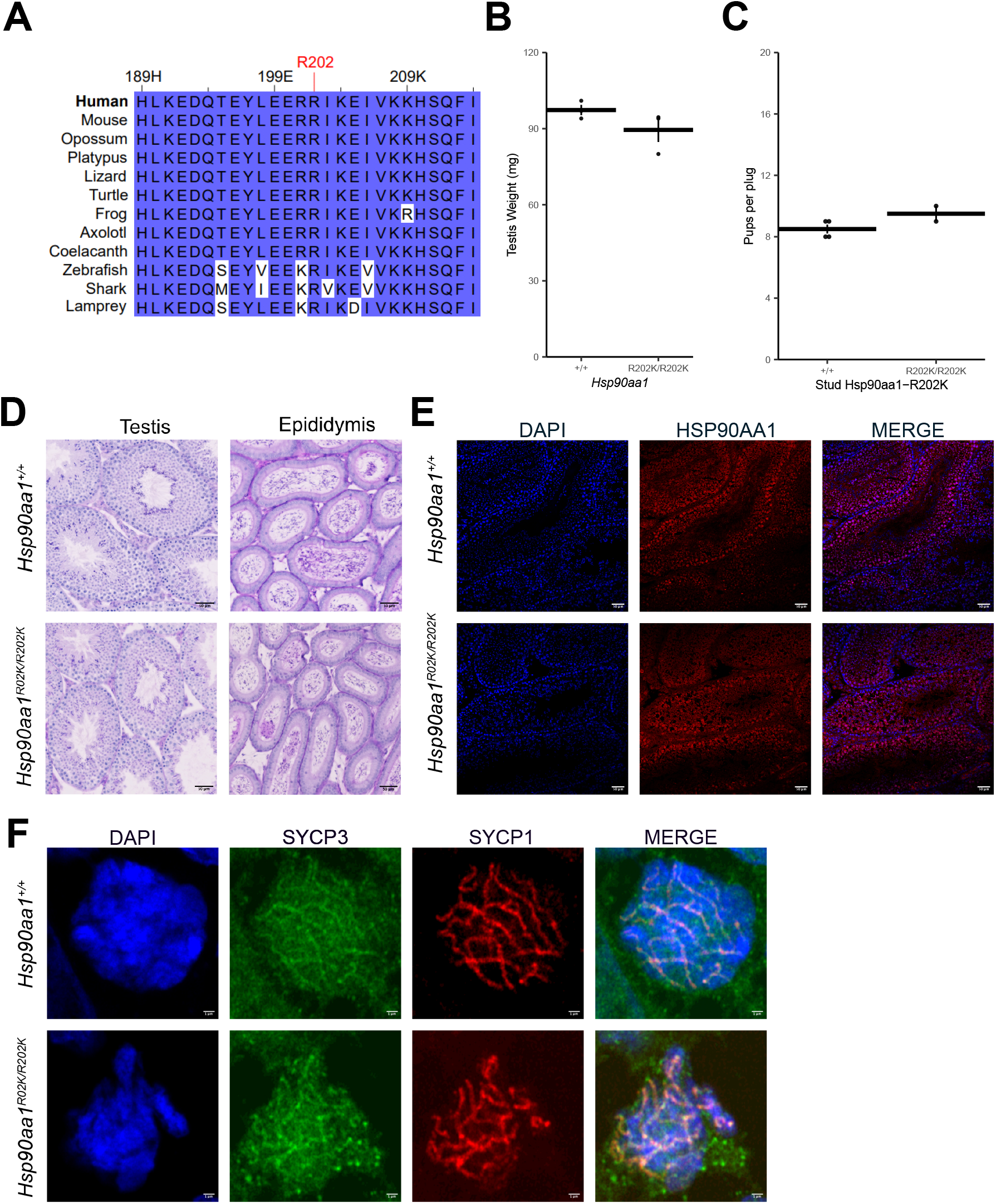
**A** The amino acid affect by the variant p.(Arg324Lys) corresponding to R202K in mice, is highly conserved. **B** Testicular weight is normal in *Hsp90aa1*^*R202K/R202K*^ mice. **C** Fertility is normal in male *Hsp90aa1*^*R202K/R202K*^ mice. **D** Testicular and epididymis histology of *Hsp90aa1*^*R202K/R202K*^ mice shows no differences compared to *Hsp90aa1*^*+/+*^ mice. **E** Expression of HSP90AA1 is not affected by the R202K variant in mice. **F** Staining for the meiotic markers SYCP1 and SYCP3 did not detect any differences between *Hsp90aa1*^*R202K/R202K*^ and *Hsp90aa1*^*+/+*^

In a first step, we assessed testicular weight and fertility of *Hsp90aa1*^*R202K/R202K*^ mice using plug matings. This analysis did not reveal a significant difference in testicular weight (Figure 1B) or the number of offspring conceived by *Hsp90aa1*^*R202K/R202K*^ mice when compared to *Hsp90aa1*^*+/+*^ mice (Figure 1C). Male and female pups were born in mendelian numbers. This finding was supported by the testicular histology, showing no difference between *Hsp90aa1*^*R202K/R202K*^ and *Hsp90aa1*^*+/+*^ mice concerning the histology of the seminiferous tubules, presence of germ cells of all stages and mature spermatozoa found in the epididymis (Figure 1D). When staining for HSP90AA1 in mouse testicular tissue, we found no difference in the protein expression (Figure 1E). Next, we focused on spermatocytes undergoing meiosis and stained for the synaptonemal complex proteins SCP1 and SCP3. We found no defects in synaptonemal complex assembly in spermatocytes from *Hsp90aa1*^*R202K/R202K*^ mice with no difference to germ cells from *Hsp90aa1*^*+/+*^ litter mates (Figure 1F). Based on our observations we concluded that the variant p.(Arg202Lys) is likely benign and not the cause of M3272’s infertility.

When going back to our sequencing data, there were no other high impact biallelic variants. Further, in gnomAD database there are no individuals with homozygous loss-of-function (LoF) variants in *HSP90AA1*.

### Heterozygous *HSP90AA1* variants

In addition to homozygous variants, we screened the exome sequencing data for heterozygous high impact variants, resulting in the detection of the frameshift variant c.1068_1071del p.(Lys357ArgfsTer20) in M1551. This man was diagnosed with hypospermatogenesis (Figure 2A) with most cells arresting at spermatocyte stage and only rare presence of post-meiotic cells (Figure 2B). This variant is located in exon 6 of 11, likely leading to a nonsense mediated decay (Figure 3A).

**Figure 2.**
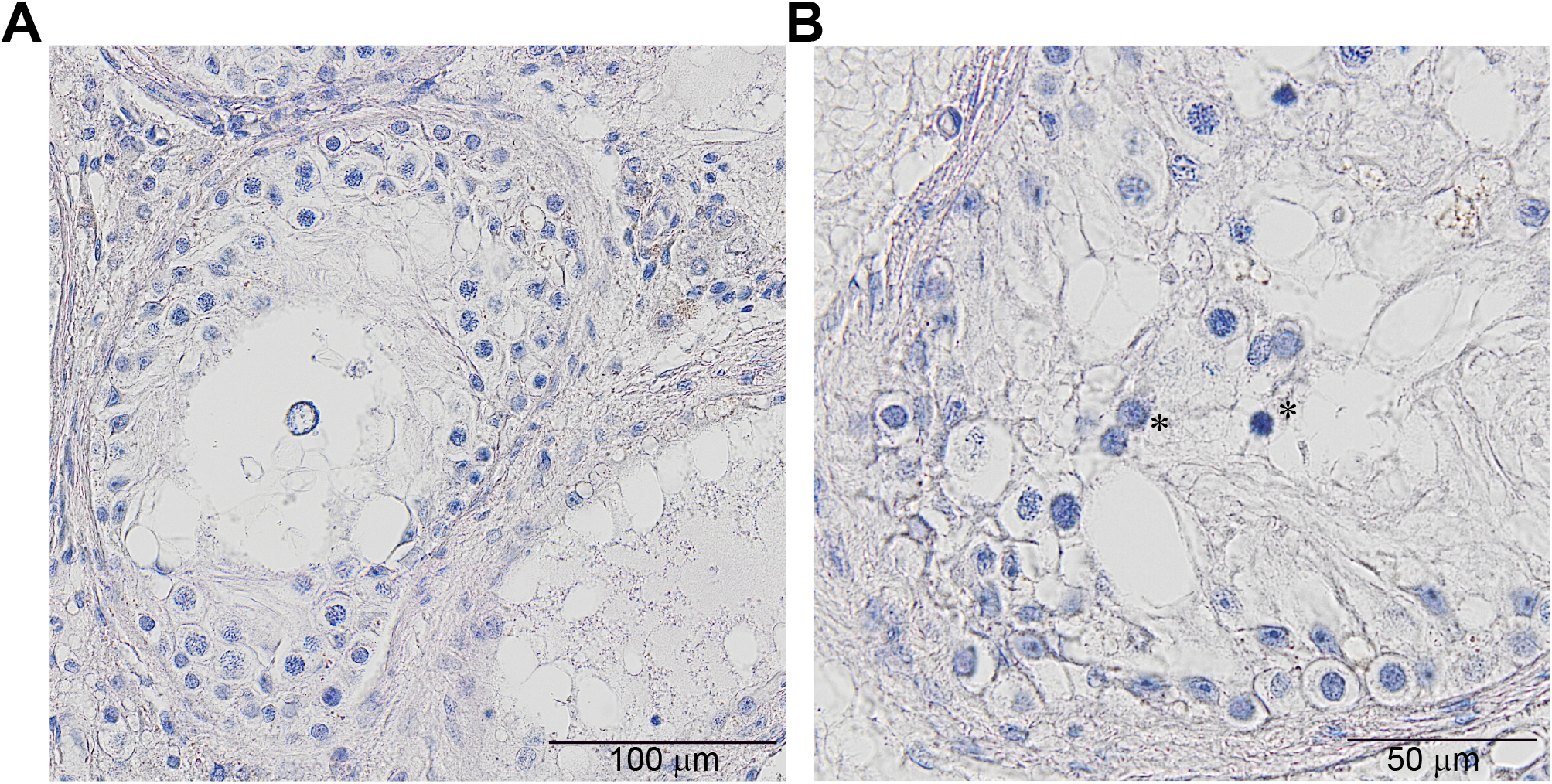
**A** Testicular histology of M1551 harbouring the heterozygous frameshift variant p.(Lys479ArgfsTer20) shows overall hypospermatogenesis with most tubules exhibiting maturation arrest at spermatocyte stage. **B** Rarely testicular tubules of M1551 show post-meiotic cells with some round spermatids (marked with *).

**Figure 3.**
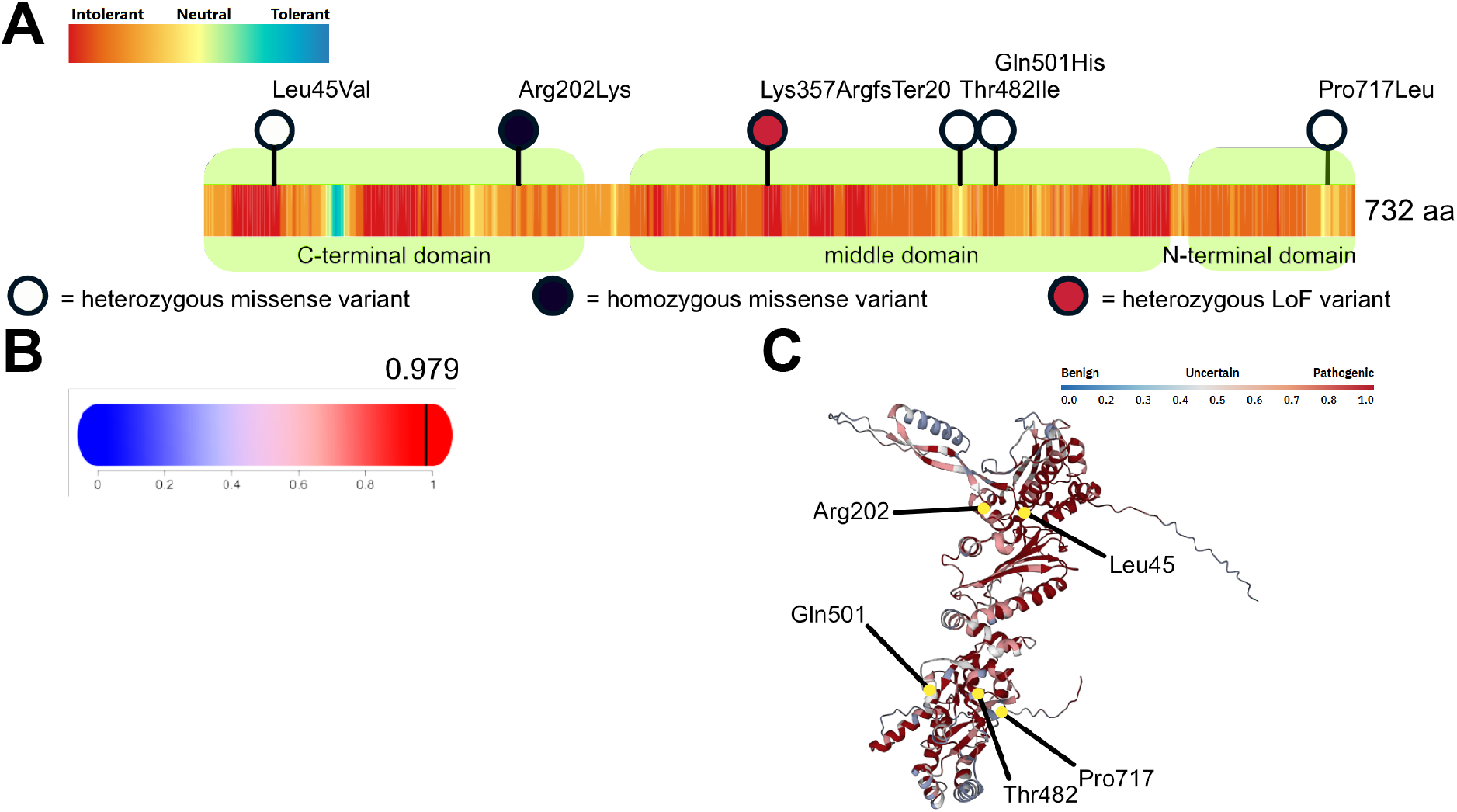
**A** Schematic overview of human HSP90AA1 and predicted landscape of tolerance to change according to Metadome with identified LoF and missense variants with a GADD score >20, MAF <0.001 and occurence <5 in our in-house database. Protein domains were taken from Haase *et al*. (PMID: 26358502). **B** Predicted autosomal dominant mode of inheritance for *HSP90AA 1* according to DOMINO. of detected missense variants are marked. **C** HSP90AA1 structure and pathogenicity of all possible human missense variants as predicted by AlphaMissense. The sites

We also identified the missense variant c.133C>G p.(Leu45Val) in M3510, which is located in the N-terminal and therefore possibly impairing ATP binding. We further found the missense variants c.1445C>T p.(Thr482Ile) in M2503 and c.1503G>C p.(Gln501His) in M2011, which are both located in the middle domain, suggesting altered client binding of the chaperone. Additionally, we identified the missense variant c.2150C>T p.(Pro717Leu) in M2643, located in the N-terminal and potentially hampering dimerization of HSP90AA1 with another HSP90AA1 protein or with HSP90AB1 (Figure 3A, 3C). The phenotypes of all these men are highly homogeneous as M2643 and M2503 have cryptozoospermia and M2011, M3510 and M1551 have azoospermia due to hypospermatogenesis, demonstrating that haploinsufficiency leads to impaired spermatogenesis but still allows formation of single elongating spermatids. These observations match the expression of HSP90AA1 with strong expression seen in spermatogonia and spermatocytes (Figure 1E). We predicted the effect of the identified missense variants on HSP90AA1 structure using AlphaMissense with the results indicating that these variants except for c.1503G>C p.(Gln501His) might be pathogenic. As we did not perform functional studies to test the effect of these variants, their functional impact remains currently unknown.

No alternative genetic causes of infertility were found when screening the exome sequencing data of men with *HSP90AA1* variants.

## Discussion

Male infertility due to reduced sperm count was shown to be of genetic origin in a substantial fraction of cases as revealed during the last ten years (Houston *et al*., 2021; James *et al*., 2025). Meanwhile more than 500 genes have been described associated with spermatogenic failure in humans (James *et al*., 2025). However, still the majority of azoospermic men remain without a causal diagnosis (Riera-Escamilla *et al*., 2025), giving rise to the assumption that there are more than the so far described genes involved in the pathogenicity of NOA. Variants in ∼75% of genes associated with spermatogenic failure in humans, follow an autosomal recessive mode of inheritance and only ∼9% of genes are autosomal dominant genes, while the percentage of dominant genes in mouse models is even smaller (James *et al*., 2025). It is therefore not unusual to find different modes of inheritance between humans and mice (Raymond *et al*., 2000; Emich *et al*., 2023).

With *HSP90AA1* we provide a novel candidate gene for spermatogenic failure, that possibly follows an autosomal dominant mode of inheritance. Although it was previously described that the single nucleotide polymorphism rs11457523 is associated with male infertility, there are no reports so far presenting variants in *HSP90AA1* as a monogenic cause of human male infertility. The fact that we did not detect any individual with biallelic LoF variants in the MERGE cohort or in gnomAD database, might implicate that humans with biallelic LoF variants in *HSP90AA1* are not viable, similar to what it is observed in *Hsp90ab1*^*-/-*^ mice. Although this hypothesis is highly speculative, we clearly noted that there seems to be a strong selection against LoF variants in *HSP90AA1* as these are extremely rare.

While HSP90AA1 is strongly expressed in human spermatogonia and spermatocytes (Thul *et al*., 2017), its exact function during spermatogenesis is unknown. Nevertheless, it is known that in mice the androgen receptor (AR) is chaperoned by HSP90AA1 in spermatogonia before its association with testosterone (Whitesell and Cook, 1996; Zhang *et al*., 2006) and that HSP90AA1 deficient mice lack expression of the AR in spermatogonia (Kajiwara *et al*., 2012). The absence of HSP90AA1 subsequently leads to a complete arrest of spermatogenesis and apoptosis of cells after pachytene stage (Kajiwara *et al*., 2012). In humans, *AR* is a well-established infertility gene with (likely) pathogenic variants in *AR* resulting in NOA with or without broader signs of androgen insensitivity (Gaikwad *et al*., 2025).

Under the assumption that the identified heterozygous variants in *HSP90AA1* are indeed the cause for the men’s infertility, there is a slight difference in the testicular phenotype observed in *Hsp90ab1*^*-/-*^ mice and affected humans. While the phenotype in mice is meiotic arrest, all men with identified variants in this study show formation of very few elongating spermatids, resulting in either hypospermatogenesis or cryptozoospermia. In men with a heterozygous variant, this might be due to a residual function of the one unaffected gene copy of *HSP90AA1*, which might still be able to provide some chaperone protein to stabilise small amounts of AR in male germ cells.

Despite these findings HSP90AA1 should not routinely be analysed in infertile men at this point, because it currently reaches a level of only limited evidence according to the ClinGen classification since we did only detect one individual with a LoF variant. Further, the mode of inheritance still remains unclear as our assessment only relies on prediction tools. To further investigate this, segregation analyses would be helpful. Unfortunately, this was not possible for the variants we detected. Detection of a *de novo* variant in *HSP90AA1* would strengthen evidence for our hypothesis. Future studies in independent patient cohorts are needed to further investigate variants in this candidate gene.

In summary, we present *HSP90AA1* as a novel candidate gene for human spermatogenic failure. It might be one of the few genes in this context, that follows an autosomal dominant mode of inheritance. If these findings can be replicated and validated in future studies, they may help narrowing the diagnostic gap in crypto- and azoospermic patients, ultimately providing more patients with a diagnosis and improving our understanding of human spermatogenesis.

**Table 1.** Identified patients with prioritised variants in *HSP90AA1*.

| Patient | Variant | Geno-<br>type | Alpha<br>Missense<br>prediction | Semen<br>phenotype | Testicular<br>histology<sup>1</sup> |
| --- | --- | --- | --- | --- | --- |
| M3510 | c.133C>G<br>p.(Leu45Val) | Hetero-<br>zygous | Likely<br>pathogenic | Azoospermia | Hypospermato-<br>genesis,<br>ES 82%/7%,<br>RS 2%/1%,<br>SC 10%/12%,<br>SG 3%/7%,<br>SCO 0%/3%,<br>TS 2%/69% |
| M3272 | c.605G>A<br>p.(Arg202Lys)<sup>2</sup> | Homo-<br>zygous | Likely<br>benign | Azoospermia | SCO |
| M1551 | c.1068_1071del<br>p.(Lys357Argfs<br>Ter20) | Hetero-<br>zygous | NA | Azoospermia | Hypospermato-<br>genesis<br>ES 0%/1%,<br>RS 7%/4%,<br>SC 48%/15%,<br>SG 2%/8%,<br>SCO 11%/35%,<br>TS 33%/37% |
| M2503 | c.1445C>T<br>p.(Thr482Ile) | Hetero-<br>zygous | ambiguous | Crypto-<br>zoospermia | unknown |
| M2011 | c.1503G>C<br>p.(Gln501His) | Hetero-<br>zygous | Likely<br>benign | Azoospermia | Hypospermato-<br>genesis,<br>no details available |
| M2643 | c.2150C>T<br>p.(Pro717Leu) | Hetero-<br>zygous | Likely<br>pathogenic | Crypto-<br>zoospermia | unknown |

<sup>1</sup>The percentages refer to the fraction of seminiferous tubules containing elongated spermatids (ES), round spermatids (RS), spermatocytes (SC), spermatogonia (SG), Sertoli cells (SCO) or tubular shadows (TS) as the most differentiated cell type/phenotype. Biopsies from the right and the left testis were analysed, resulting in two different numbers per cell type/phenotype per patient.
<sup>2</sup>Variant likely benign due to fertile mouse model.

## Data Availability

All data produced in the present study are available upon reasonable request to the authors.

## Acknowledgements

The authors are grateful to all individuals for having agreed to use their DNA, testicular samples and data for research use. The authors are grateful for the technical assistance and support provided by Luisa Meier. The authors thank David Kelly at the Centre for Optical Instrumentation Laboratory (COIL) and the Shared University Research Facilities (SURF) histology and histological imaging facility for support with histology of mouse samples.

This project was funded by a German Research Foundation (DFG) fellowship (award WY 215/1-1 to MJW), the DFG-sponsored Clinical Research Unit ‘Male Germ Cells’ (CRU326, project 329621271 to FT), and Wellcome Trust funding (225237 to DOC). This work was supported by funding for the Wellcome Discovery Research Platform for Hidden Cell Biology (226791).

---

**Figure S1.**
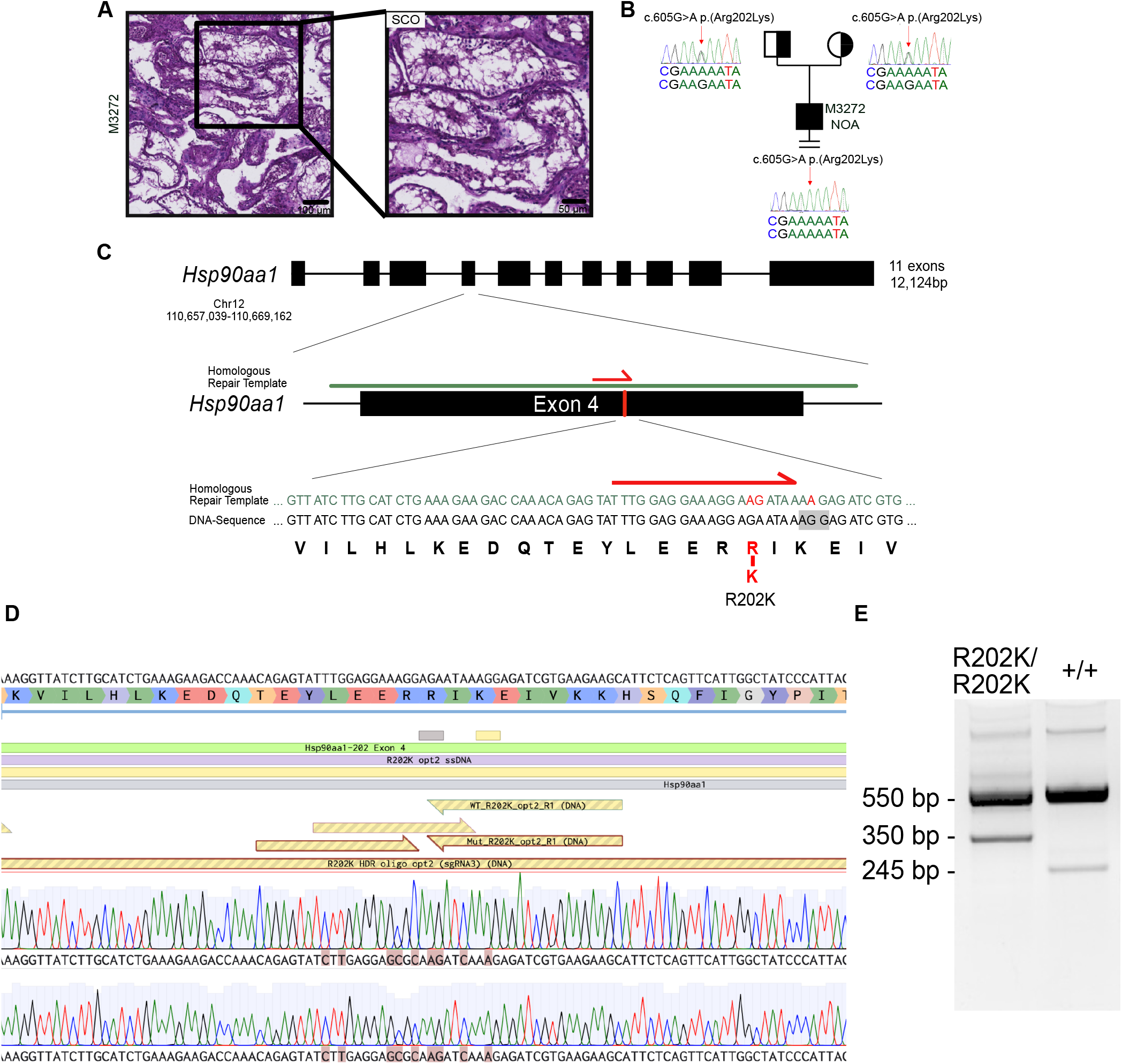
**A** Testicular histology of M3272 showing SCO. **B** Segregation analysis of the missense variant c.605G>A p.(Arg202Lys) showed that M3272 is homozygous for this variant while both parents are heterozygous carriers. **C** Strategy for targeting *Hsp90aa1* and creation of the R202K allele in mice. **E** PCR genotyping of *Hsp90aa1*^*R202K/R202K*^ and *Hsp90aa1*^*+/+*^ mice. **D** Sequencing of *Hsp90aa1*^*R202K/R202K*^ mice confirms homozygosity for this allele.

